# Differences in Skilled Nursing Facility Characteristics and Quality Ratings by Long-Term Care Pharmacy Provider

**DOI:** 10.1101/2021.08.31.21262894

**Authors:** Andrew R. Zullo, Melissa R. Riester, Elizabeth M. Goldberg, Meghan A. Cupp, Sarah D. Berry, Francesca L. Beaudoin

## Abstract

**Objectives:** Limited data exist on the U.S. long-term care (LTC) pharmacy market and how skilled nursing facilities (SNFs) may differ by LTC pharmacy provider. We estimated the market shares of two major LTC pharmacies (Omnicare and PharMerica) and assessed if SNF characteristics differ by pharmacy.

**Design:** Cross-sectional.

**Setting and Participants:** Seventy-five Rhode Island (RI) SNFs that provided post-acute care (PAC) services in 2019.

**Methods:** SNF location, structure, staffing measures, and quality ratings were ascertained from publicly available data sources. The LTC pharmacy used by each SNF was compiled by case managers at a RI health system.

**Results:** Among 75 SNFs, 32 (43%) were served by Omnicare and 36 (48%) by PharMerica. LASSO logistic regression and random forest models identified 5 key predictors of SNFs selecting PharMerica over Omnicare: number of skilled beds, total number of beds, nursing hours per resident per day, five-star health inspection rating, and average number of residents per day. In a multivariable regression model including 4 predictors (total number of beds excluded due to collinearity), SNFs had a 6% higher prevalence of using Omnicare over PharMerica for every additional 10 skilled beds (Prevalence Ratio 1.06, 95% CI 1.02-1.10).

**Conclusions and Implications:** Omnicare and PharMerica comprised over 90% of the SNF PAC market in RI, with Omnicare covering larger facilities. Understanding if these companies serve a similar proportion of SNFs in other U.S. states is necessary to advance future research initiatives and examine how collaborations between SNFs and LTC pharmacy chains may improve medication management in SNFs.

**Brief Summary:** Two pharmacies may comprise over 90% of the skilled nursing facility post-acute care market. This market concentration may represent a prime opportunity to efficiently improve medication management.

## INTRODUCTION

As demand for post-acute care (PAC) continues to increase in the United States, long-term care (LTC) pharmacies play a progressively greater role in skilled nursing facility (SNF) care through the provision of prescription medications, medication management, and other services. Due to evolving payment systems and increasing financial pressure, LTC pharmacies have consolidated through mergers and acquisitions over recent years.^1^ This has resulted in two companies dominating the U.S. market—Omnicare Inc. (a wholly owned subsidiary of CVS Health Corporation) and PharMerica Corporation. Although their exact combined market share is unknown, Omnicare and PharMerica may now provide a majority of prescription drugs to U.S. SNFs.^1-3^

The predominance of two LTC pharmacies offers a unique opportunity for policymakers, payors, healthcare executives, researchers, and other healthcare stakeholders to improve medication management and SNF quality of care through collaborative interventions between SNFs and the two LTC pharmacy partners. New programs and clinical pharmacy services could be delivered by the LTC pharmacies to the thousands of SNFs they serve, effectively intervening on a majority of U.S. SNFs. Such programs to optimize medication management are particularly important for SNF residents since polypharmacy is common and an estimated 40% of individuals in SNFs take ≥9 medications.^4-8^

The types of SNFs each LTC pharmacy chain serves and the clinical services provided to each facility (beyond medication dispensing) are currently unknown. In the absence of data on how SNFs differ by LTC pharmacy provider, including facility characteristics (e.g., SNF size) and quality ratings, researchers and policymakers are limited in their ability to design collaborative interventions or policies that improve medication management in SNFs. Further, without such information, interventions cannot be tailored to the attributes and unique needs of the SNFs served by each pharmacy.

Our objectives were to 1) provide empirical estimates of the market shares of each major LTC pharmacy (Omnicare and PharMerica), and 2) assess if SNFs’ structure, staffing, and quality characteristics differ by LTC pharmacy provider. We hypothesized that, based on prior public communications from Omnicare and PharMerica to their investors, 1) Omnicare would serve a larger proportion of SNFs than PharMerica, and 2) the SNFs served by Omnicare would be larger facilities with higher overall five-star quality ratings.

## METHODS

### Study Design and Data Sources

This cross-sectional study linked publicly available SNF-level data from calendar year 2019 to private data on the LTC pharmacies used by each SNF in the state of Rhode Island (RI). The Nursing Home Compare website and RI Department of Health Nursing Home Summary Report provided SNF-level information on structure, staffing, and quality for each facility in the state.^9, 10^ Since obtaining accurate information on which LTC pharmacy a SNF uses is challenging, we used RI as an exemplar because we could leverage existing partnerships to accurately ascertain that information. The LTC pharmacy for each SNF was collected by the case managers of a local health system in December 2019. These case managers refer patients to SNFs to receive PAC after an inpatient hospital stay and coordinate care transitions by ensuring that discharge prescriptions are sent to the correct LTC pharmacy. Case managers created the LTC pharmacy list as part of their routine workflow and updated the list at regular intervals. Since 1) a single LTC pharmacy is typically used for all patients receiving PAC within the same SNF and 2) the LTC pharmacy list is essential for operations, all case managers confirmed that data were highly accurate. To verify, the lead investigator (A.R.Z.) called 10 randomly sampled SNFs and spoke with the SNF staff to independently confirm their LTC pharmacy provider. No inaccuracies were identified. All datasets were SNF-level; therefore, this study did not require Institutional Review Board approval.

### Study Population

To maximize generalizability, we included all RI SNFs that provided PAC. Assisted living facilities, facilities that provided only LTC (no PAC), and those without SNF beds were excluded (n= 95).

### LTC Pharmacies of Interest

Our primary interest was whether a SNF contracted with Omnicare versus PharMerica as its LTC pharmacy. We defined this dependent variable in analyses as SNF use of Omnicare versus PharMerica.

### SNF Characteristics

SNF characteristics (enumerated in Supplementary Figure S1) were selected based on their potential to influence the use of Omnicare over PharMerica. Selection of characteristics was informed by statements made by Omnicare and PharMerica to investors and the public, complaints documented by the Federal Trade Commission, and discussions with the case managers at the local health system.^1-3^ SNF characteristics of interest included location, structure, capacity, occupancy, staffing, and five-star quality ratings.

### Statistical Analysis

We used descriptive statistics to summarize the frequency and prevalence of Omnicare versus PharMerica use among SNFs, then described SNF characteristics overall and stratified by LTC pharmacy provider. To understand how SNF characteristics were associated with use of Omnicare versus PharMerica, we used t-tests for continuous variables and chi-squared tests for categorical variables (or their exact or non-parametric equivalents whenever necessary). To understand which SNF characteristics might be the most important predictors of using one LTC pharmacy over another, we used least absolute shrinkage and selection operator (LASSO) logistic regression models with adaptive selection to select SNF characteristics.^11, 12^ In a stability analysis to test the robustness of our primary LASSO approach, we used random forests with 4-fold cross-validation to identify key predictors, then generated corresponding variable importance plots. Finally, after checking correlations between the key SNF characteristic predictors identified through the LASSO logistic regression and random forest, we specified a final multivariable modified Poisson regression model with robust standard errors to estimate prevalence ratios (PRs) with 95% confidence intervals (95% CIs).^13^

## RESULTS

### LTC Pharmacy SNF Market Share

Of the 75 RI SNFs that provided PAC services in 2019, 68 (91%) were served by Omnicare or PharMerica. Thirty-two of the 75 SNFs (43%) were served by Omnicare and 36 (48%) were served by PharMerica. Among the SNFs not served by Omnicare or PharMerica, 4 (5%) were served by ProCare LTC, a LTC pharmacy chain serving New England and Mid-Atlantic states, and 3 (4%) were served by White Cross Pharmacy, a LTC pharmacy serving RI and southeastern Massachusetts.

### Characteristics of SNFs by LTC Pharmacy

Overall, the 75 SNFs in the study sample had a median (first quartile, third quartile) of 100 (60, 133) total beds, 31 (18, 54) skilled beds, and 91 (55, 124) residents per day (Table 1). Compared to SNFs served by Omnicare, SNFs served by PharMerica had fewer total beds (median, 90 versus 124), fewer skilled beds (median, 20 versus 34), and fewer residents per day (median, 75 versus 105). There were no differences in five-star ratings between SNFs using PharMerica versus Omnicare in bivariable analyses (Table 2). Characteristics for SNFs served by other LTC pharmacies are included in Supplemental Digital Content 1.

**Table 1.** Characteristics of Skilled Nursing Facilities Overall and Stratified by Long-Term Care Pharmacy Provider, 2019.

| Characteristic | Overall<sup>a</sup> | Omnicare | PharMerica | P-value |
| --- | --- | --- | --- | --- |
| Facility N | 75 | 32 | 36 |  |
| County |  |  |  | 0.36 |
| Providence | 43 (57) | 19 (59) | 21 (58) |  |
| Kent | 11 (15) | 5 (16) | 6 (17) |  |
| Bristol | 5 (7) | 3 (9) | 0 (0) |  |
| Washington | 10 (13) | 4 (13) | 6 (17) |  |
| Newport | 6 (8) | 1 (3) | 3 (8) |  |
| Beds, All, no., median (Q1, Q3) | 100 (60, 133) | 124 (61, 149) | 90 (60, 120) | 0.12 |
| Beds, All, no., mean (SD) | 106 (51) | 116 (58) | 96 (45) | 0.12 |
| Beds, Skilled, no., median (Q1, Q3) | 31 (18, 54) | 34 (22, 57) | 20 (15, 43) | 0.01 |
| Beds, Skilled, no., mean (SD) | 43 (37) | 52 (43) | 31 (24) | 0.01 |
| Average Number of Residents / Day, mean (SD) | 95 (47) | 101 (52) | 87 (42) | 0.24 |
| Average Number of Residents / Day, median (IQR) | 91 (55, 124) | 105 (48, 128) | 75 (56, 114) | 0.32 |
| Nursing Hours / Resident / Day, mean (SD) | 0.84 (0.23) | 0.83 (0.23) | 0.88 (0.22) | 0.32 |
| Nursing Hours / Resident / Day, median (IQR) | 0.88 (0.65, 1.03) | 0.89 (0.64, 1.03) | 0.88 (0.75, 1.03) | 0.51 |
| For-Profit Status | 15 (20) | 5 (16) | 9 (25) | 0.34 |
| Medicare-certified | 75 (100) | 32 (100) | 36 (100) | 1.00 |
| Medicaid-certified | 75 (100) | 32 (100) | 36 (100) | 1.00 |
| Secure Dementia Care Unit | 30 (40) | 11 (34) | 16 (44) | 0.40 |
| *Abbreviations:* Q1, first quartile; Q3, third quartile; SD, standard deviation.<br>*Notes:* Characteristics reported as number (percent [%]), unless otherwise specified. <sup>a</sup>Includes the 7 skilled nursing facilities that were served by long-term care pharmacies other than Omnicare and PharMerica. Supplemental Digital Content 1 includes characteristics for skilled nursing facilities served by other long-term care pharmacies. |  |  |  |  |

**Table 2.**
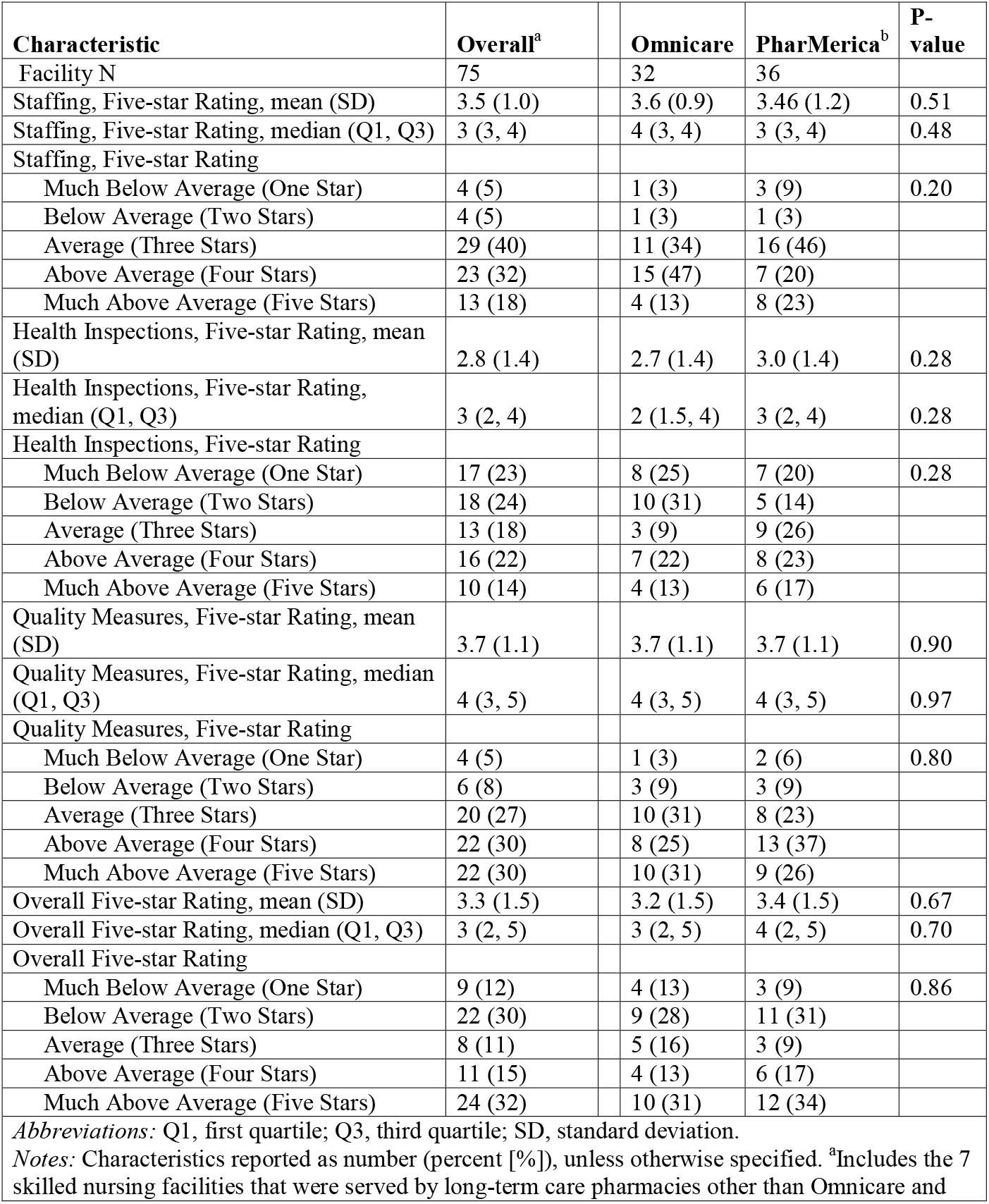
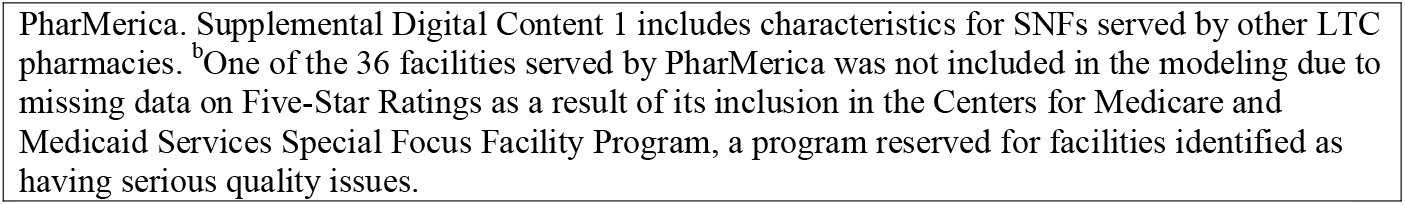
Five-Star Quality Ratings of Skilled Nursing Facilities Overall and Stratified by Long-Term Care Pharmacy Provider, 2019.

| Characteristic | Overall<sup>a</sup> | Omnicare | PharMerica<sup>b</sup> | P-value |
| --- | --- | --- | --- | --- |
| Facility N | 75 | 32 | 36 |  |
| Staffing, Five-star Rating, mean (SD) | 3.5 (1.0) | 3.6 (0.9) | 3.46 (1.2) | 0.51 |
| Staffing, Five-star Rating, median (Q1, Q3) | 3 (3, 4) | 4 (3, 4) | 3 (3, 4) | 0.48 |
| Staffing, Five-star Rating |  |  |  |  |
| Much Below Average (One Star) | 4 (5) | 1 (3) | 3 (9) | 0.20 |
| Below Average (Two Stars) | 4 (5) | 1 (3) | 1 (3) |  |
| Average (Three Stars) | 29 (40) | 11 (34) | 16 (46) |  |
| Above Average (Four Stars) | 23 (32) | 15 (47) | 7 (20) |  |
| Much Above Average (Five Stars) | 13 (18) | 4 (13) | 8 (23) |  |
| Health Inspections, Five-star Rating, mean (SD) | 2.8 (1.4) | 2.7 (1.4) | 3.0 (1.4) | 0.28 |
| Health Inspections, Five-star Rating, median (Q1, Q3) | 3 (2, 4) | 2 (1.5, 4) | 3 (2, 4) | 0.28 |
| Health Inspections, Five-star Rating |  |  |  |  |
| Much Below Average (One Star) | 17 (23) | 8 (25) | 7 (20) | 0.28 |
| Below Average (Two Stars) | 18 (24) | 10 (31) | 5 (14) |  |
| Average (Three Stars) | 13 (18) | 3 (9) | 9 (26) |  |
| Above Average (Four Stars) | 16 (22) | 7 (22) | 8 (23) |  |
| Much Above Average (Five Stars) | 10 (14) | 4 (13) | 6 (17) |  |
| Quality Measures, Five-star Rating, mean (SD) | 3.7 (1.1) | 3.7 (1.1) | 3.7 (1.1) | 0.90 |
| Quality Measures, Five-star Rating, median (Q1, Q3) | 4 (3, 5) | 4 (3, 5) | 4 (3, 5) | 0.97 |
| Quality Measures, Five-star Rating |  |  |  |  |
| Much Below Average (One Star) | 4 (5) | 1 (3) | 2 (6) | 0.80 |
| Below Average (Two Stars) | 6 (8) | 3 (9) | 3 (9) |  |
| Average (Three Stars) | 20 (27) | 10 (31) | 8 (23) |  |
| Above Average (Four Stars) | 22 (30) | 8 (25) | 13 (37) |  |
| Much Above Average (Five Stars) | 22 (30) | 10 (31) | 9 (26) |  |
| Overall Five-star Rating, mean (SD) | 3.3 (1.5) | 3.2 (1.5) | 3.4 (1.5) | 0.67 |
| Overall Five-star Rating, median (Q1, Q3) | 3 (2, 5) | 3 (2, 5) | 4 (2, 5) | 0.70 |
| Overall Five-star Rating |  |  |  |  |
| Much Below Average (One Star) | 9 (12) | 4 (13) | 3 (9) | 0.86 |
| Below Average (Two Stars) | 22 (30) | 9 (28) | 11 (31) |  |
| Average (Three Stars) | 8 (11) | 5 (16) | 3 (9) |  |
| Above Average (Four Stars) | 11 (15) | 4 (13) | 6 (17) |  |
| Much Above Average (Five Stars) | 24 (32) | 10 (31) | 12 (34) |  |
| *Abbreviations:* Q1, first quartile; Q3, third quartile; SD, standard deviation. |  |  |  |  |
| *Notes:* Characteristics reported as number (percent [%]), unless otherwise specified. <sup>a</sup>Includes the 7 skilled nursing facilities that were served by long-term care pharmacies other than Omnicare and |  |  |  |  |

PharMerica. Supplemental Digital Content 1 includes characteristics for SNFs served by other LTC pharmacies. <sup>b</sup>One of the 36 facilities served by PharMerica was not included in the modeling due to missing data on Five-Star Ratings as a result of its inclusion in the Centers for Medicare and Medicaid Services Special Focus Facility Program, a program reserved for facilities identified as having serious quality issues.

### Identifying Predictors of SNFs Utilizing Omnicare versus PharMerica

The LASSO logit identified two key predictors of SNFs selecting Omnicare versus PharMerica: number of skilled beds and health inspection five-star rating. Results from the stability analysis using random forests in the full set (Supplementary Figure S1) of variables were partially consistent with those from the LASSO; the variables with the highest importance scores included number of skilled beds, average number of residents per day, total number of beds, and nursing hours per resident per day.

### Key Predictors of SNFs Utilizing Omnicare versus PharMerica

The final multivariable Poisson model included the union of the two sets of variables identified from the LASSO logit and random forest: number of skilled beds (per 10 bed increase), average number of residents per day (per 10 resident increase), nursing hours per resident per day (per 1-hour increase), and five-star health inspection rating (≥3 stars versus <3 stars). Total number of beds was not advanced because it was highly collinear with average number of residents per day (Pearson correlation coefficient= 0.97). In the multivariable model, SNFs had a 6% higher prevalence of using Omnicare versus PharMerica for every additional 10 skilled beds in the SNF (PR 1.06, 95% CI 1.02-1.10) (Table 3). No other remarkable differences were observed.

**Table 3.** Multivariable Associations Between Skilled Nursing Facility Characteristics and Use of Omnicare versus PharMerica as the Long-Term Care Pharmacy, 2019 (N=67^a^).

| Characteristic | Multivariable Prevalence Ratio (95% CI), p-value |
| --- | --- |
| Skilled Beds (per 10 bed increase) | 1.06 (1.02-1.10), p=0.001 |
| Average Number of Residents / Day (per 10 resident increase) | 1.01 (0.95-1.08), p=0.707 |
| Nursing Hours / Resident / Day (per 1-hour increase) | 1.38 (0.33-5.71), p=0.658 |
| Health Inspection Five-Star Rating |  |
| One or two stars | 1.00 (reference) |
| Three or more stars | 0.61 (0.36-1.04), p=0.070 |
| *Abbreviations:* CI, confidence interval.<br>*Notes:* <sup>a</sup>One of the 36 facilities served by PharMerica was not included in the modeling due to missing data on Five-Star Ratings as a result of its inclusion in the Centers for Medicare and Medicaid Services Special Focus Facility Program, a program reserved for facilities identified as having serious quality issues. |  |

## DISCUSSION

In this cross-sectional study of 75 SNFs offering PAC in RI, 32 (43%) SNFs were served by Omnicare and 36 (48%) were served by PharMerica, indicating that these large LTC pharmacy chains comprised 91% of the RI SNF PAC market in 2019. Facilities using Omnicare tended to be larger on average, but SNF staffing and quality ratings did not differ significantly by LTC pharmacy provider. If these companies also serve a majority of SNFs across the U.S., a significant opportunity may exist to improve medication management and SNF care quality on a large scale through partnerships between SNFs and these predominant LTC pharmacies.

Interventions delivered pragmatically through LTC pharmacies could improve patient care and clinical outcomes for SNF residents and may also result in added benefits to the SNF in terms of quality measures and cost savings.^14^ For example, interventions to monitor antipsychotic prescribing for residents with behavioral symptoms of Alzheimer’s disease and related dementias (ADRD) could promote gradual dose reductions and eventual discontinuation of antipsychotic medications, thereby reducing the risk of potentially severe medication adverse effects.^15^ SNFs that successfully deprescribe might also improve their five-star quality rating since antipsychotic use measures are reported through the Nursing Home Compare website and the Five-Star Quality Rating System.^16, 17^ LTC pharmacies may also be ideally positioned to assist with transitions of care between hospitals and SNFs to reduce potentially avoidable hospital readmissions, because many rehospitalizations are medication-related.^18, 19^ Reducing potentially avoidable readmissions is increasingly important since CMS regulations now incentivize SNFs to make quality improvements based on performance on hospital readmission measures.^20^

Understanding how SNF characteristics vary by LTC pharmacy providers and predictors of SNFs selecting one LTC pharmacy provider over another are crucial precursor steps to developing and implementing effective medication management programs that can be embedded in dominant LTC pharmacy chains. Future research programs may wish to customize quality improvement initiatives based on whether the LTC pharmacy chain serves facilities with a greater number of beds or lower quality ratings. To inform which programs and services may be most effective for LTC pharmacies to implement, future research should first examine which clinical pharmacy services are currently available to SNFs (e.g., consultant pharmacist services, programs targeted at certain conditions such as ADRD), how often these services are utilized by SNFs, who provides these services (e.g., LTC pharmacy, pharmacist employed by the SNF), and if the services improve medication-related outcomes and are cost-effective.

Finally, understanding the characteristics and predictors of SNFs served by each LTC pharmacy will help address a variety of policy issues. For example, understanding which SNFs each LTC pharmacy serves could inform discussions on new payment and care delivery models for PAC. Additionally, it may be important to examine how continued horizontal and vertical consolidation of LTC pharmacies and SNFs switching from one pharmacy to another affects patient outcomes and Medicare expenditures.^2^

### Limitations

Our study has at least two key limitations. First, since our study included only RI SNFs, our results may not generalize to the entire U.S. SNF PAC market. The parent company of Omnicare, CVS Health Corporation, is headquartered in RI, which could potentially have impacted Omnicare’s market share in the state. Second, our statistical power was limited due to our modest sample size. Despite these limitations, our results provide one of the first estimates of the combined market share of Omnicare and PharMerica and predictors of SNFs using one LTC pharmacy chain over another.

## CONCLUSIONS AND IMPLICATIONS

This study examined the LTC pharmacy market share in a single state and assessed how SNF characteristics differ by LTC pharmacy provider. Our findings demonstrated that two LTC pharmacy chains dominated the SNF PAC market. The U.S. LTC pharmacy market and the types of clinical services that LTC pharmacies deliver to SNFs beyond medication dispensing should be a focus of future inquiry. Understanding national differences in SNF characteristics by LTC pharmacy provider will help policymakers, payors, researchers, and others to begin to understand the current contribution of LTC pharmacies to SNF quality of care and the potential role for interventions to improve SNF quality related to medications.

## Supporting information

Supplemental Digital Content 1

## Data Availability

Data are available upon reasonable request from the authors.

## Acknowledgements

The authors thank Dr. Vincent Mor for his feedback on early versions of the manuscript.

---

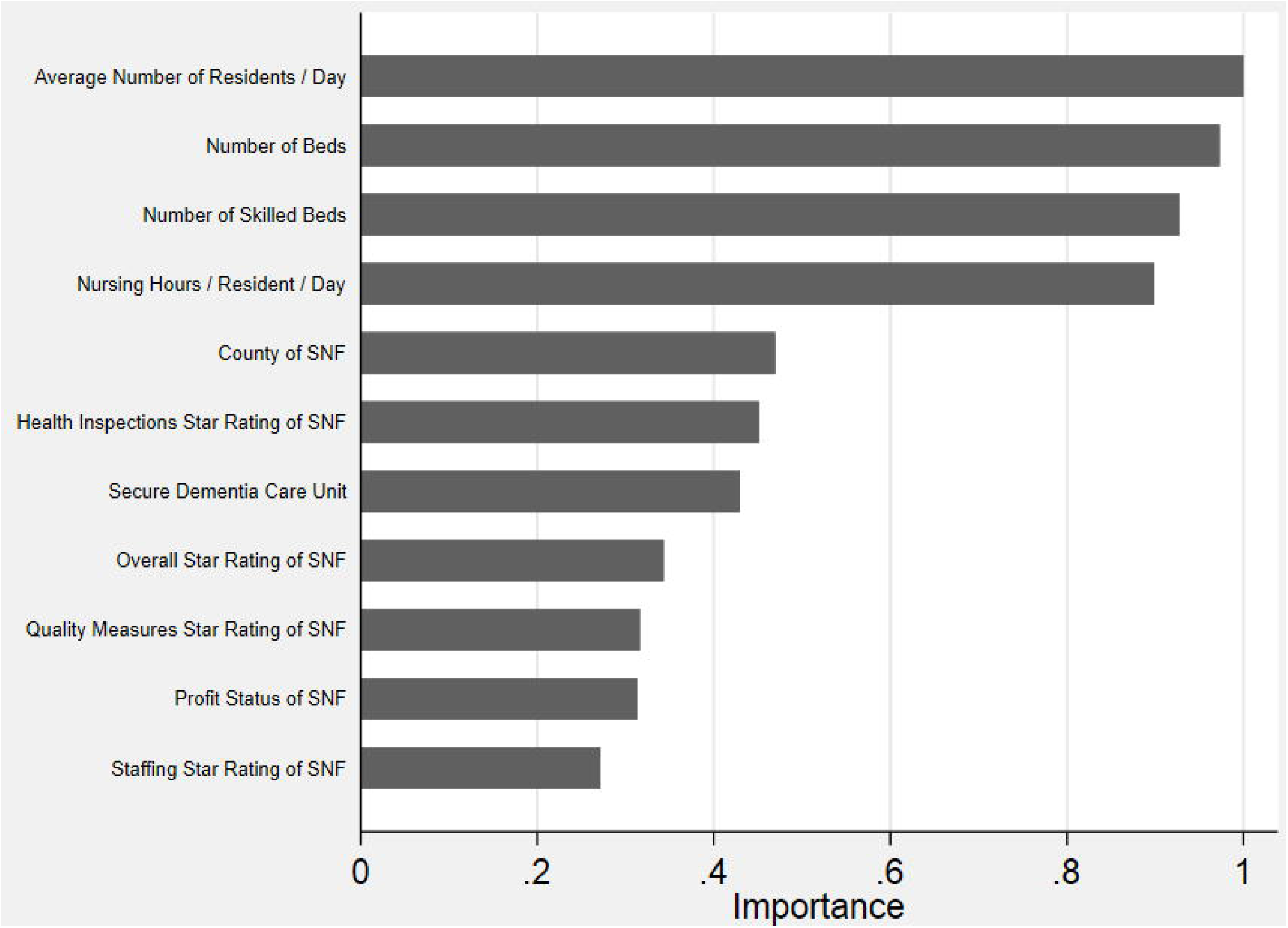

## REFERENCES

1. Oliver K. Active immunity: The aging population will strengthen industry revenue. IBISWorld Industry Report OD5707 Institutional Pharmacies in the US; 2018.

2. Avalere. Long-Term Care Pharmacy: the Evolving Marketplace and Emerging Policy Issues. 2015.

3. In the Matter of Omnicare, Inc. a corporation, United States of America Before the Federal Trade Commission, (2012).

4. Dwyer L, Han B, Woodwell D, Ea R. Polypharmacy in nursing home residents in the United States: results of the 2004 National Nursing Home Survey. Am J Geriatr Pharmacother. 2010;

5. Zullo AR, Smith RJ, Gutman R, et al. Comparative safety of dipeptidyl peptidase-4 inhibitors and sulfonylureas among frail older adults. Journal of the American Geriatrics Society. Jul 21 2021;doi:10.1111/jgs.17371

6. Zullo AR, Lee Y, Lary C, Daiello LA, Kiel DP, Berry SD. Comparative effectiveness of denosumab, teriparatide, and zoledronic acid among frail older adults: a retrospective cohort study. Osteoporosis international : a journal established as result of cooperation between the European Foundation for Osteoporosis and the National Osteoporosis Foundation of the USA. Mar 2021;32(3):565–573. doi:10.1007/s00198-020-05732-2

7. Zullo AR, Riester MR, Erqou S, Wu WC, Rudolph JL, Steinman MA. Comparative Effectiveness of Angiotensin II Receptor Blockers and Angiotensin-Converting Enzyme Inhibitors in Older Nursing Home Residents After Myocardial Infarction: A Retrospective Cohort Study. Drugs & aging. Oct 2020;37(10):755–766. doi:10.1007/s40266-020-00791-w

8. Zullo AR, Ofori-Asenso R, Wood M, et al. Effects of Statins for Secondary Prevention on Functioning and Other Outcomes Among Nursing Home Residents. Journal of the American Medical Directors Association. Apr 2020;21(4):500–507 e8. doi:10.1016/j.jamda.2020.01.102

9. Medicare.gov. Find & compare nursing homes, hospitals & other providers near you. Accessed July 9, 2021, https://www.medicare.gov/care-compare/

10. State of Rhode Island Department of Health. Nursing Home Quality. Accessed July 9, 2021, https://health.ri.gov/nursinghomes/about/quality/

11. Buhlmann P, van de Geer S. Statistics for High-Dimensional Data: Methods, Theory and Applications. Springer; 2011.

12. Zou H. The Adaptive Lasso and Its Oracle Properties. J Am Stat Assoc. 2006;101(476):1418–1429.

13. Zou G. A modified poisson regression approach to prospective studies with binary data. American journal of epidemiology. Apr 1 2004;159(7):702–6. doi:10.1093/aje/kwh090

14. MHA. Procopio R. The Value of the Long-Term Care Pharmacist in the Delivery and Continuum of Care. Accessed July 29, 2021, https://www.mhainc.com/uploadedFiles/Content/Resources/MHA%20The%20Value%20of%20the%20LTC%20Pharmacist%200720%20-%20final.pdf

15. Levinson D. Medicare atypical antipsychotic drug claims for elderly nursing home residents. Department of Health and Human Services Office of Inspector General; 2011.

16. Gurwitz J, Bonner A, Berwick D. Reducing Excessive Use of Antipsychotic Agents in Nursing Homes. JAMA. 2017;318(2):118–119.

17. Centers for Medicare & Medicaid Services. Design for Nursing Home Compare Five-Star Quality Rating System: Technical Users’ Guide.

18. Boockvar K, Fishman E, Kyriacou C, Monias A, Gavi S, Cortes T. Adverse events due to discontinuations in drug use and dose changes in patients transferred between acute and longterm care facilities.. Arch Intern Med. 2004;164(5):545–550.

19. Boockvar K, Liu S, Goldstein N, Nebeker J, Siu A, Fried T. Prescribing discrepancies likely to cause adverse drug events after patient transfer. Qual Saf Health Care. 2009;18(1):32–36.

20. 80 FR 46389 Medicare Program; Prospective Payment System and Consolidated Billing for Skilled Nursing Facilities (SNFs) for FY 2016, SNF Value-Based Purchasing Program, SNF Quality Reporting Program, and Staffing Data Collection 46389-46477 (2015).

