## Supplemental Digital Content 1 for "Differences in Skilled Nursing Facility Characteristics and Quality Ratings by Long-Term Care Pharmacy Provider"

**Affiliations:** ^a^Department of Health Services, Policy, and Practice, Brown University School of Public Health, 121 South Main Street, Providence, RI, USA; ^b^Department of Epidemiology, Brown University School of Public Health, 121 South Main Street, Providence, RI, USA; ^c^Center of Innovation in Long-Term Services and Supports, Providence Veterans Affairs Medical Center, 830 Chalkstone Ave, Building 32, Providence, RI, USA; ^d^Department of Pharmacy, Rhode Island Hospital, 593 Eddy Street, Rhode Island Hospital, Providence, RI, USA; ^e^Department of Emergency Medicine, Rhode Island Hospital, 593 Eddy Street, Rhode Island Hospital, Providence, RI, USA. ^f^Department of Medicine, 330 Brookline Avenue, Beth Israel Deaconess Medical Center, Boston, MA; ^g^Marcus Institute for Aging Research, Hebrew SeniorLife, 1200 Centre Street, Boston, MA.

**Corresponding Author:** Andrew R. Zullo, PharmD, PhD, Department of Health Services, Policy, and Practice, Brown University School of Public Health, 121 South Main Street, Box G-S121-8, Providence, RI, USA, 02912, Phone: 401-863-6309,, Twitter: @andrewzullo.

**Supplemental Table S1.** Characteristics of Skilled Nursing Facilities Overall and Stratified by Long-Term Care Pharmacy Provider, 2019.

**Supplemental Table S2.** Five-Star Quality Ratings of Skilled Nursing Facilities Overall and Stratified by Long-Term Care Pharmacy Provider, 2019.

**Supplemental Figure S1.** Importance Scores for Associations between Skilled Nursing Facility Characteristics and Use of Omnicare versus PharMerica from Random Forest Stability Analysis (N=67). *The random forest model used 4-fold cross-validation (i.e., each of the 25% samples of the overall dataset were successively used as a test set while the remaining 75% of the data is used as the training set) to identify key predictors of using one LTC pharmacy over another. Note: One facility served by PharMerica was not included in the modeling due to missing data on five-star ratings.*

**Supplemental Table 1.** Characteristics of Skilled Nursing Facilities Overall and Stratified by Long-Term Care Pharmacy Provider, 2019.

| **Characteristic** | **Overall** | **Omnicare** | **PharMerica** | **Other LTC Pharmacy** |
| --- | --- | --- | --- | --- |
| Facility N | 75 | 32 | 36 | 7 |
| County |  |  |  |  |
| Providence | 43 (57) | 19 (59) | 21 (58) | 3 (43) |
| Kent | 11 (15) | 5 (16) | 6 (17) | 0 (0) |
| Bristol | 5 (7) | 3 (9) | 0 (0) | 2 (29) |
| Washington | 10 (13) | 4 (13) | 6 (17) | 0 (0) |
| Newport | 6 (8) | 1 (3) | 3 (8) | 2 (29) |
| Beds, All, no., median (Q1, Q3) | 100 (60, 133) | 124 (61, 149) | 90 (60, 120) | 114 (76, 166) |
| Beds, All, no., mean (SD) | 106 (51) | 116 (58) | 96 (45) | 113 (47) |
| Beds, Skilled, no., median (Q1, Q3) | 31 (18, 54) | 34 (22, 57) | 20 (15, 43) | 46 (32, 76) |
| Beds, Skilled, no., mean (SD) | 43 (37) | 52 (43) | 31 (24) | 61 (53) |
| Average no. of residents per day, mean (SD) | 95 (47) | 101 (52) | 87 (42) | 107 (41) |
| Average no. of residents per day, median (Q1, Q3) | 91 (55, 124) | 105 (48, 128) | 75 (56, 114) | 106 (65, 156) |
| Nursing Hours / Resident / Day, mean (SD) | 0.84 (0.23) | 0.83 (0.23) | 0.88 (0.22) | 0.66 (0.22) |
| Nursing Hours / Resident / Day, median (Q1, Q3) | 0.88 (0.65, 1.03) | 0.89 (0.64, 1.03) | 0.88 (0.75, 1.03) | 0.57 (0.5, 0.9) |
| For-Profit Status | 15 (20) | 5 (16) | 9 (25) | 1 (14) |
| Medicare-certified | 75 (100) | 32 (100) | 36 (100) | 7 (100) |
| Medicaid-certified | 75 (100) | 32 (100) | 36 (100) | 7 (100) |
| Secure Dementia Care Unit | 30 (40) | 11 (34) | 16 (44) | 3 (43) |
| *Abbreviations:* LTC, long-term care pharmacy; Q1, first quartile; Q3, third quartile; SD, standard deviation.  *Notes:* Characteristics reported as number (percent [%]), unless otherwise specified. | | | | |

**Supplemental Table 2.** Five-Star Quality Ratings of Skilled Nursing Facilities Overall and Stratified by Long-Term Care Pharmacy Provider, 2019.

| **Characteristic** | **Overall** | **Omnicare** | **PharMerica**^a^ | **Other LTC Pharmacy^b^** |
| --- | --- | --- | --- | --- |
| Facility N | 75 | 32 | 36 | 7 |
| Staffing, Five-star Rating, mean (SD) | 3.5 (1.0) | 3.6 (0.9) | 3.5 (1.1) | 3.2 (1.2) |
| Staffing, Five-star Rating, median (Q1, Q3) | 3 (3, 4) | 4 (3, 4) | 3 (3, 4) | 3 (2, 4) |
| Staffing, Five-star Rating |  |  |  |  |
| Much Below Average (One Star) | 4 (5) | 1 (3) | 3 (9) | 0 (0) |
| Below Average (Two Stars) | 4 (5) | 1 (3) | 1 (3) | 2 (33) |
| Average (Three Stars) | 29 (40) | 11 (34) | 16 (46) | 2 (33) |
| Above Average (Four Stars) | 23 (32) | 15 (47) | 7 (20) | 1 (17) |
| Much Above Average (Five Stars) | 13 (18) | 4 (13) | 8 (23) | 1 (17) |
| Health Inspections, Five-star Rating, mean (SD) | 2.8 (1.4) | 2.7 (1.4) | 3.0 (1.4) | 2.1 (1.1) |
| Health Inspections, Five-star Rating, median (Q1, Q3) | 3 (2, 4) | 2 (2, 4) | 3 (2, 4) | 2 (1, 3) |
| Health Inspections, Five-star Rating |  |  |  |  |
| Much Below Average (One Star) | 17 (23) | 8 (25) | 7 (20) | 2 (29) |
| Below Average (Two Stars) | 18 (24) | 10 (31) | 5 (14) | 3 (43) |
| Average (Three Stars) | 13 (18) | 3 (9) | 9 (26) | 1 (14) |
| Above Average (Four Stars) | 16 (22) | 7 (22) | 8 (23) | 1 (14) |
| Much Above Average (Five Stars) | 10 (14) | 4 (13) | 6 (17) | 0 (0) |
| Quality Measures, Five-star Rating, mean (SD) | 3.7 (1.1) | 3.7 (1.1) | 3.7 (1.1) | 3.7 (1.5) |
| Quality Measures, Five-star Rating, median (Q1, Q3) | 4 (3, 5) | 4 (3, 5) | 4 (3, 5) | 4 (3, 5) |
| Quality Measures, Five-star Rating |  |  |  |  |
| Much Below Average (One Star) | 4 (5) | 1 (3) | 2 (6) | 1 (14) |
| Below Average (Two Stars) | 6 (8) | 3 (9) | 3 (9) | 0 (0) |
| Average (Three Stars) | 20 (27) | 10 (31) | 8 (23) | 2 (29) |
| Above Average (Four Stars) | 22 (30) | 8 (25) | 13 (37) | 1 (14) |
| Much Above Average (Five Stars) | 22 (30) | 10 (31) | 9 (26) | 3 (43) |
| Overall Five-star Rating, mean (SD) | 3.3 (1.5) | 3.2 (1.5) | 3.4 (1.5) | 2.9 (1.8) |
| Overall Five-star Rating, median (Q1, Q3) | 3 (2, 5) | 3 (2, 5) | 4 (2, 5) | 2 (1, 5) |
| Overall Five-star Rating |  |  |  |  |
| Much Below Average (One Star) | 9 (12) | 4 (13) | 3 (9) | 2 (29) |
| Below Average (Two Stars) | 22 (30) | 9 (28) | 11 (31) | 2 (29) |
| Average (Three Stars) | 8 (11) | 5 (16) | 3 (9) | 0 (0) |
| Above Average (Four Stars) | 11 (15) | 4 (13) | 6 (17) | 1 (14) |
| Much Above Average (Five Stars) | 24 (32) | 10 (31) | 12 (34) | 2 (29) |
| *Abbreviations:* Q1, first quartile; Q3, third quartile; SD, standard deviation.  *Notes:* Characteristics reported as number (percent (%)), unless otherwise specified.  ^a^One facility had missing data on all five-star ratings due to inclusion in the Centers for Medicare and Medicaid Services Special Focus Facility Program, a program reserved for facilities identified as having serious quality issues.  ^b^One facility had insufficient data to obtain an adequate staffing five-star rating. | | | | |

**Supplemental Figure S1.** Importance Scores for Associations between Skilled Nursing Facility Characteristics and Use of Omnicare versus PharMerica from Random Forest Stability Analysis (N=67).

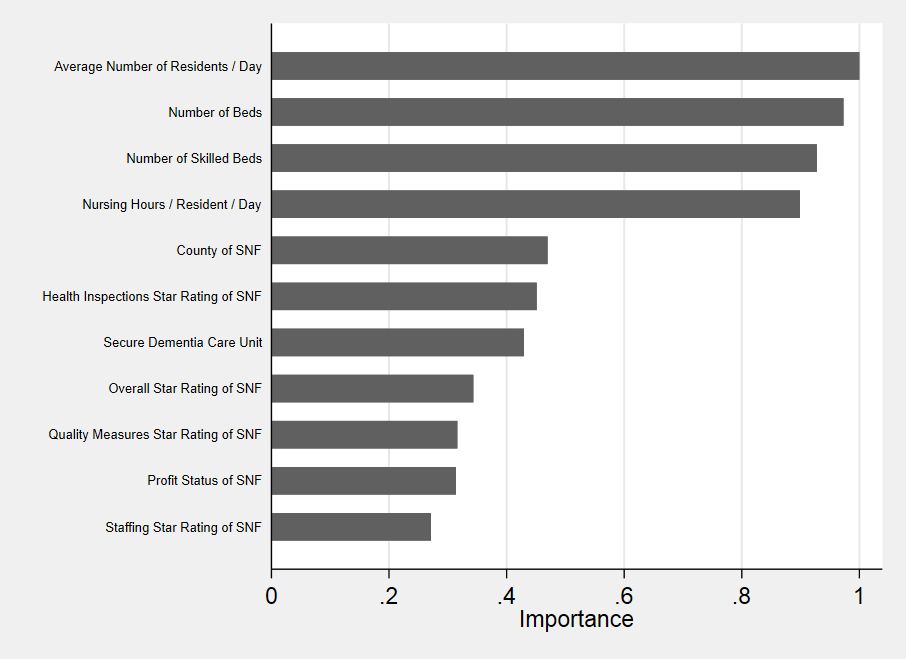

*The random forest model used 4-fold cross-validation (i.e., each of the 25% samples of the overall dataset were successively used as a test set while the remaining 75% of the data is used as the training set) to identify key predictors of using one LTC pharmacy over another. Note: One facility served by PharMerica was not included in the modeling due to missing data on five-star ratings.*
